# Assessment of the laboratory capacity for testing Sexual Transmitted Infections at 14 health facilities in Moshi Municipality, Tanzania

**DOI:** 10.64898/2026.05.13.26353104

**Authors:** Victor V. Mosha, Emanuel Samky, Gloria Ngowi, Martha Msemwa, Deborah Macha, Winfrida Mwita, Werner Maokola, Johnson Lyimo, Odile B. Harrison, Sia E. Msuya

**Author notes:** Corresponding author: Victor Mosha. These authors contributed equally to this work (Odile B. Harrison, Sia E. Msuya).

## Abstract

The global occurrence of sexually transmitted infections (STIs) continues to rise, necessitating accurate diagnosis and treatment to curb their spread and associated complications. With the alarming increase in antimicrobial resistance (AMR) in Neisseria gonorrhoeae, effective STI management relies heavily on etiological diagnosis. The Tanzania National Standard for Medical Laboratories 2017 outlines recommended STI testing protocols based on facility levels, yet adherence to these guidelines and associated challenges remain poorly documented. This study describes the diagnostic capacity for different STIs in northern Tanzania. A cross-sectional study was conducted between May and July 2023, encompassing 14 laboratories across Moshi Municipal Council, Kilimanjaro region. The laboratories assessed were in five hospitals and nine health centres (HCs). Data regarding facility type and STI diagnostic capabilities were gathered through questionnaires administered during site visits and supplemented by observations. All five hospitals were equipped to conduct rapid diagnostic tests for HIV, syphilis, and wet preparation microscopy for Trichomonas vaginalis (TV). Only three hospitals had the capacity to perform culture and sensitivity testing using chocolate and blood agar medium, however none reported isolating Neisseria gonorrhoeae in the past year. Critical STI diagnostic tests including the Treponema pallidum particle agglutination assay (TPPA) and Treponema pallidum hemagglutination assay (TPHA) for the laboratory confirmation of syphilis, assays for Chlamydia trachomatis, Herpes Simplex virus –2, and Human papillomavirus (HPV) were absent across all five hospitals. Conversely, all health centers demonstrated proficiency in rapid treponemal tests for syphilis, together with rapid HIV test and TV testing, although one health center lacked the capacity for wet laboratory preparation for TV detection. Findings underscore a concerning lack of STI testing capacity within surveyed healthcare facilities, posing significant barriers to effective STI management and exacerbating the threat of AMR in Tanzania. In particular, the capacity for conventional microbiology culture was limited in most settings, severely compromising the ability to track and monitor AMR. Urgent investment in laboratory infrastructure and training is imperative to enhance STI diagnosis and treatment, ultimately curtailing transmission and mitigating the impact of AMR.

## Introduction

Sexually transmitted infections (STIs) are primarily spread through sexual contact. They are caused by multiple pathogens, including bacteria, viruses, and parasites, and are transmitted through bodily fluids or skin-to-skin contact during vaginal, oral, or anal sex (1). Globally, more than 1 million STIs are acquired every day in individuals aged 15–49 (2). In 2020, an estimated 374 million new cases of four curable STIs: chlamydia (*Chlamydia trachomatis,* CT), gonorrhoea (*Neisseria gonorrhoeae,* Ng), syphilis (*Treponema pallidum subsp pallidum,* TP), and trichomoniasis (*Trichomonas vaginalis,* TV) were reported worldwide in 15-49 year olds, most of which from low– and middle-income countries (LMICs) (2).

STIs can spread upward through the female urogenital tract, resulting in conditions such as pelvic and tubal inflammation that compromise reproductive health and increase the risk of infertility, ectopic pregnancy, miscarriage, and other adverse pregnancy outcomes (3). These infections can also lead to neonatal complications, including conjunctivitis, sepsis, and meningitis (4). The emergence and spread of AMR (5) and the lack of effective vaccines, particularly for bacterial STIs (6), limit our ability to treat these infections effectively. Improved diagnostic capacity and technical expertise are therefore critically needed, especially in LMICs where the burden of STIs is highest (7).

However, the true STI burden in low-resource settings may be much higher due to limited diagnostic tools and expertise, combined with syndromic STI management (8). Syndromic treatment has played an essential role in reducing the burden of STIs for many years (9) as it is cheap, provides instant treatment at first visit and does not require laboratory diagnostics and training (10). However, a significant limitation is that many STIs will be asymptomatic and cannot be identified without diagnostic testing (11). Syndromic management of STIs, may in addition lead to misdiagnosis, delayed treatment, and overtreatment, as the signs and symptoms of different STIs often overlap between organisms (12). For example, the syndromic management of Ng and CT infections in women is complicated as vaginal discharge may not necessarily be apparent (13). In addition, the rapid increase of AMR in Ng in different settings globally (14,15) shows the indispensable need of aetiological diagnosis in the treatment of STIs to limit inappropriate antibiotic treatment and curb AMR development.

The World Health Organisation (WHO) Global Progress Report on STI control highlights significant gaps in diagnostics and management, leading to inadequate public health responses in multiple regions worldwide. In response, the WHO has identified key research priorities across diagnostics, management, prevention, and epidemiology. In particular, the development of low-cost, rapid point-of-care (POC) tests for Ng, CT and syphilis were critical priorities, as well as tests to detect AMR in Ng and *Mycoplasma genitalium* (MG). The WHO further emphasized the need for developing multiplex diagnostic platforms to improve the detection and management of STIs (16).

In Tanzania, STIs remain a significant public health concern, and syndromic management continues to be the standard of care. As a result, most Tanzanian STI patients lack access to laboratory diagnostics. Individual studies have demonstrated that STIs persist as a problem among key populations, including young people (17), pregnant women (18,19) and the general adult population. The Ministry of Health developed the Tanzania National Standard for Medical Laboratories in 2017 to address recent advancements and achievements in medical laboratory services, focusing on supporting accreditation. The standard aims to guide the efficient use of limited resources to meet the growing demand for quality laboratory services nationwide. It provides instructions on the types of tests to be conducted at different levels of health facilities, including tests for STIs (20).

Given the WHO’s recommendation for laboratory diagnostics to enable efficient and cost-effective STI management, assessing the capacity of healthcare facilities to provide such services is critical. Here, the laboratory capacity of hospitals and primary health care facilities in Moshi municipality, located in north-eastern Tanzania were assessed. STI tests performed at these health facilities were compared against the national standards for medical laboratory requirements to identify gaps and opportunities for improvement in STI diagnostics and management.

## Materials and Methods

### Study design and setting

This was a cross-sectional study conducted between May and July 2023 in the Moshi Municipal Council, located in north-eastern Tanzania. Moshi Municipal is one of seven districts within the Kilimanjaro Region. The district has a total of 47 health facilities, including 5 hospitals, 13 health centers, and 29 dispensaries. In Tanzania, hospitals and health centers are expected to be equipped with laboratories capable of performing STI testing. The study initially aimed to include all 5 hospitals and 13 health centers within the municipality, however, not all facilities were able to participate. As a result, the study involved 14 facilities including: 5 hospitals (comprising one zonal referral hospital, one regional referral hospital, and three district-level hospitals) and the 9 health centers within the Moshi Municipal Council (Fig 1).

**Fig 1.** Map showing the locations of participating healthcare facilities in Moshi Municipal Council.

### National Standard for Medical Laboratories 2017

The Tanzania National Standard for Medical Laboratories of 2017 outlines the laboratory tests and equipment recommended for various levels of healthcare facilities, including dispensaries, health centers, and hospitals. This study focuses on the section of the standard related to STI diagnostic tests, specifically those that should be available at health centers and hospitals. Table 1 provides a detailed summary of the STI diagnostic tests recommended for implementation in health centers and hospitals as per the national guidelines.

**Table 1:**
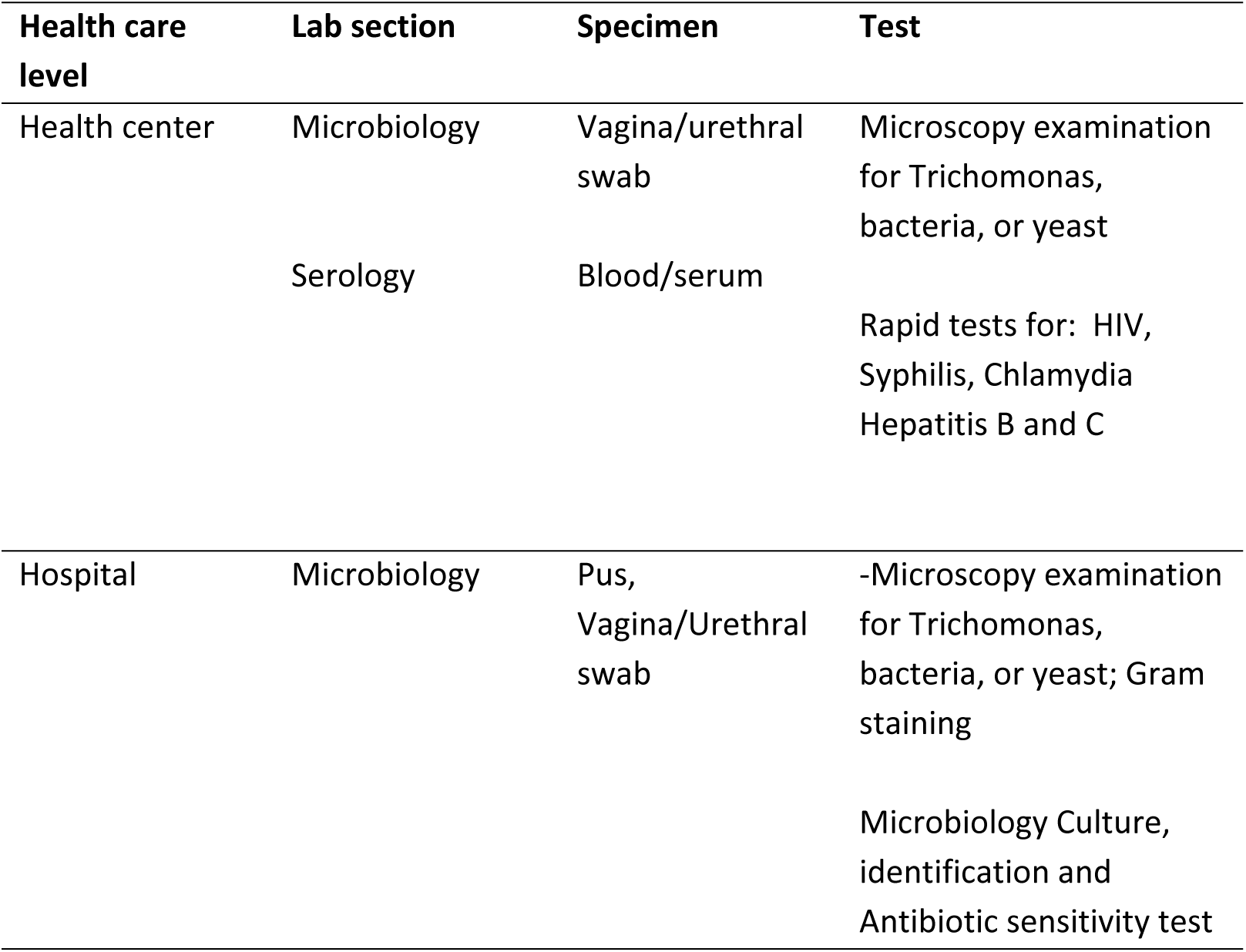
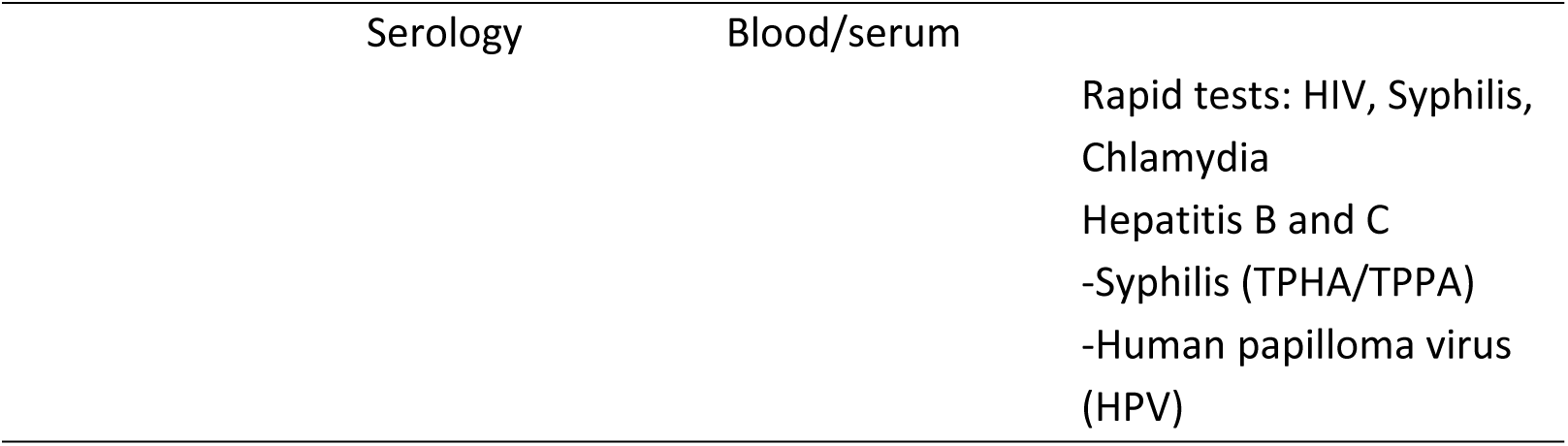
Minimum requirements for STI tests in health centers and hospitals.

### Data collection methods and procedures

Data were collected through interviews, observations, and document reviews at participating healthcare facilities. Questionnaires were used to interview representative laboratory personnel (laboratory scientists or technologist), gathering information on the health facilities, availability of STI tests, equipment, training records, and competence documented in personnel files.

The maintenance of equipment used in STI testing, such as pipettes, microscopes, refrigerators, hot plates, incubators, and autoclaves, was assessed through service reports and inspection of maintenance stickers provided by service engineers. Facilities were requested to provide Standard Operating Procedures (SOPs) covering quality control, equipment handling, operation, and maintenance.

For identified STI tests, quality control programs for reagents and associated documentation were reviewed. Verification and validation processes for methods and reagents were evaluated. In addition, SOPs for the reception, storage, acceptance testing, and inventory management of reagents and consumables were examined. Each facility’s inventory system was assessed to determine its availability and proper usage.

## Ethics approval and consent to participate

The study was approved by the Kilimanjaro Christian Medical University College Research Ethics Review Committee (CREC) under certificate number UG 32/2023. Written informed consent was also obtained from healthcare worker representatives prior to conducting interviews and observations.

### Data analysis

Data collected were entered, cleaned, and analyzed using SPSS version 23. Descriptive statistical analysis was conducted to assess the capacity of healthcare facilities in Moshi Municipality to conduct STI testing. Frequencies and percentages were used to describe categorical variables, including the availability of various STI diagnostic tests. Measures of central tendency with respective measures of dispersion were used to summarize continuous variables such as the number of STI tests performed monthly by each facility. To provide insights into adherence to national laboratory guidelines, proportions were calculated to show the percentage of facilities maintaining equipment records, conducting quality control checks, and having standard operating procedures (SOPs) in place.

## Results

### Characteristics of health facilities

Hospitals in Moshi Municipal reported a median monthly outpatient load of 8,400 (IQR: 640–10,000) and a median inpatient load of 600 (IQR: 60–900). Similarly, health centers reported a median monthly outpatient load of 750 (IQR: 660–2,930) and a median inpatient load of 100 (IQR: 70–150). For STI testing, hospitals conducted a median of 40 tests per month (range: 4–1,226), while health centers also reported a median of 40 tests per month (range: 20–100) (Table 1).

**Table 1:**
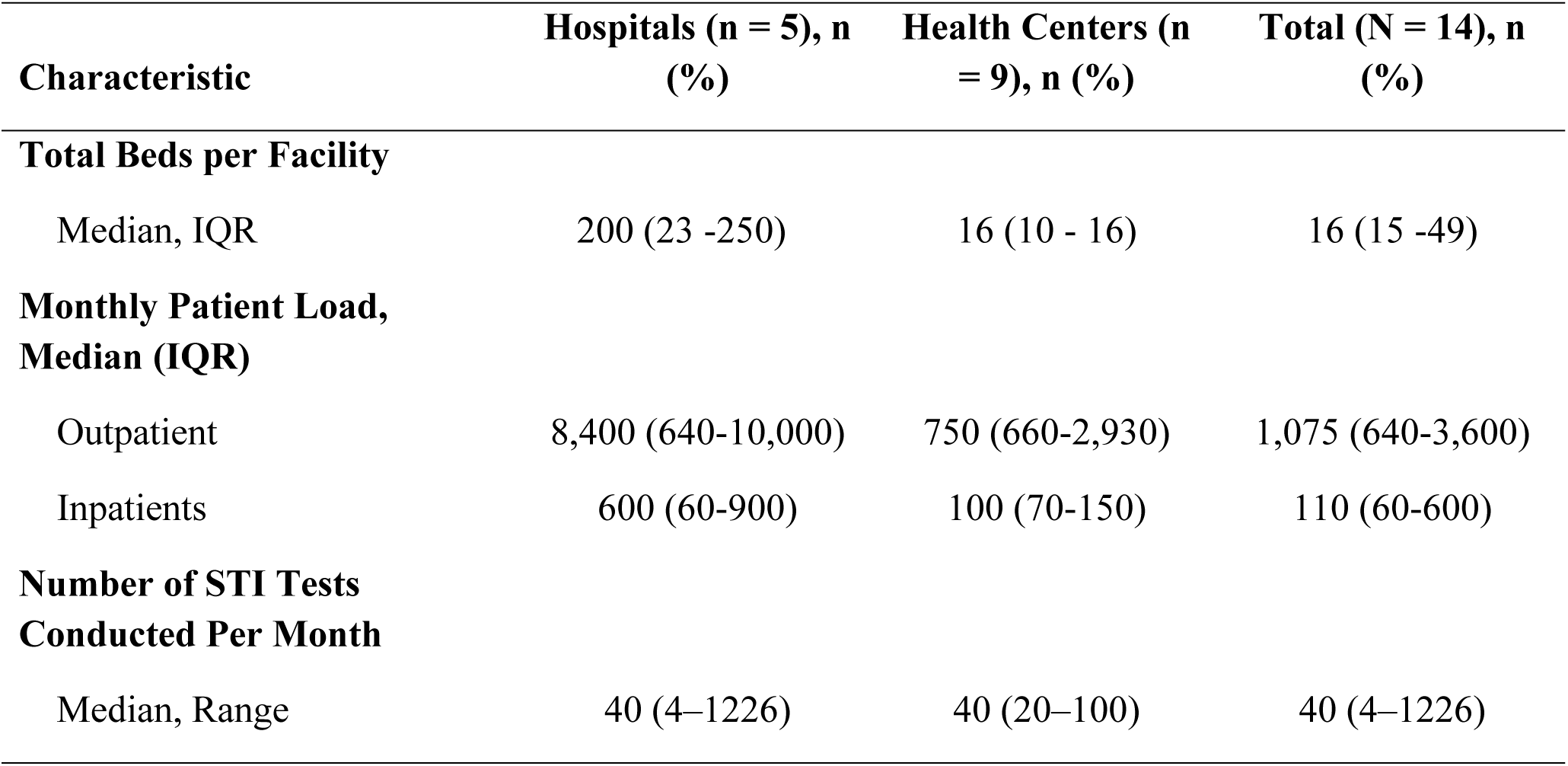
Characteristics of Health Facilities included in the study (N=14)

### Type of STIs tests available in the facilities assessed

The study revealed that all healthcare facilities, including hospitals and health centers, conducted rapid HIV and syphilis testing. Hepatitis B and C testing were performed in 80% of hospitals and 44% of health centers. Wet mount microscopy for *Trichomonas vaginalis* (TV) was widely available, being conducted in 80% of hospitals and 100% of health centers. Results also show that only 3 out of 5 hospitals had the capacity for Ng culture using chocolate agar, but none reported isolating Ng in the past year. Additionally, no facility conducted testing for CT, HPV, or laboratory confirmatory syphilis testing (TPHA/TPPA) (Table 2).

**Table 2:** Availability of STI Diagnostic Tests in Healthcare Facilities (N=14)

| STI Diagnostic Test | Hospitals (n = 5), n (%) | Health Centers (n = 9), n (%) | Total (N = 14), n (%) |
| --- | --- | --- | --- |
| HIV RDT | 5 (100) | 9 (100) | 14 (100) |
| Syphilis RDT | 5 (100) | 9 (100) | 14 (100) |
| Hepatitis B RDT | 4 (80) | 4 (44) | 8 (57) |
| Hepatitis C RDT | 4 (80) | 4 (44) | 8 (57) |
| Wet Mount for Trichomonas | 4 (80) | 9 (100) | 13 (93) |
| Culture for *Neisseria gonorrhoeae* | 3 (60) | N/A | 3 (21) |
| Chlamydia Test | 0 (0) | 0 (0) | 0 (0) |
| Syphilis TPHA/TPPA | 0 (0) | N/A | 0 (0) |
| Human Papillomavirus Test | 0 (0) | N/A | 0 (0) |

RDT: Rapid Diagnostic Tests

### Adherence to Equipment, Consumable and Reagents Standards

The assessment revealed that 71% of healthcare facilities had maintenance records for equipment, but only 29% had Standard Operating Procedures (SOPs) for equipment operation. Quality control programs for reagents were available in 71% of facilities; however, reagent validation or verification was conducted in only 14%. Additionally, only 36% of facilities had SOPs for inventory management, while 71% had an inventory system for reagents and consumables (Table 3).

**Table 3:**
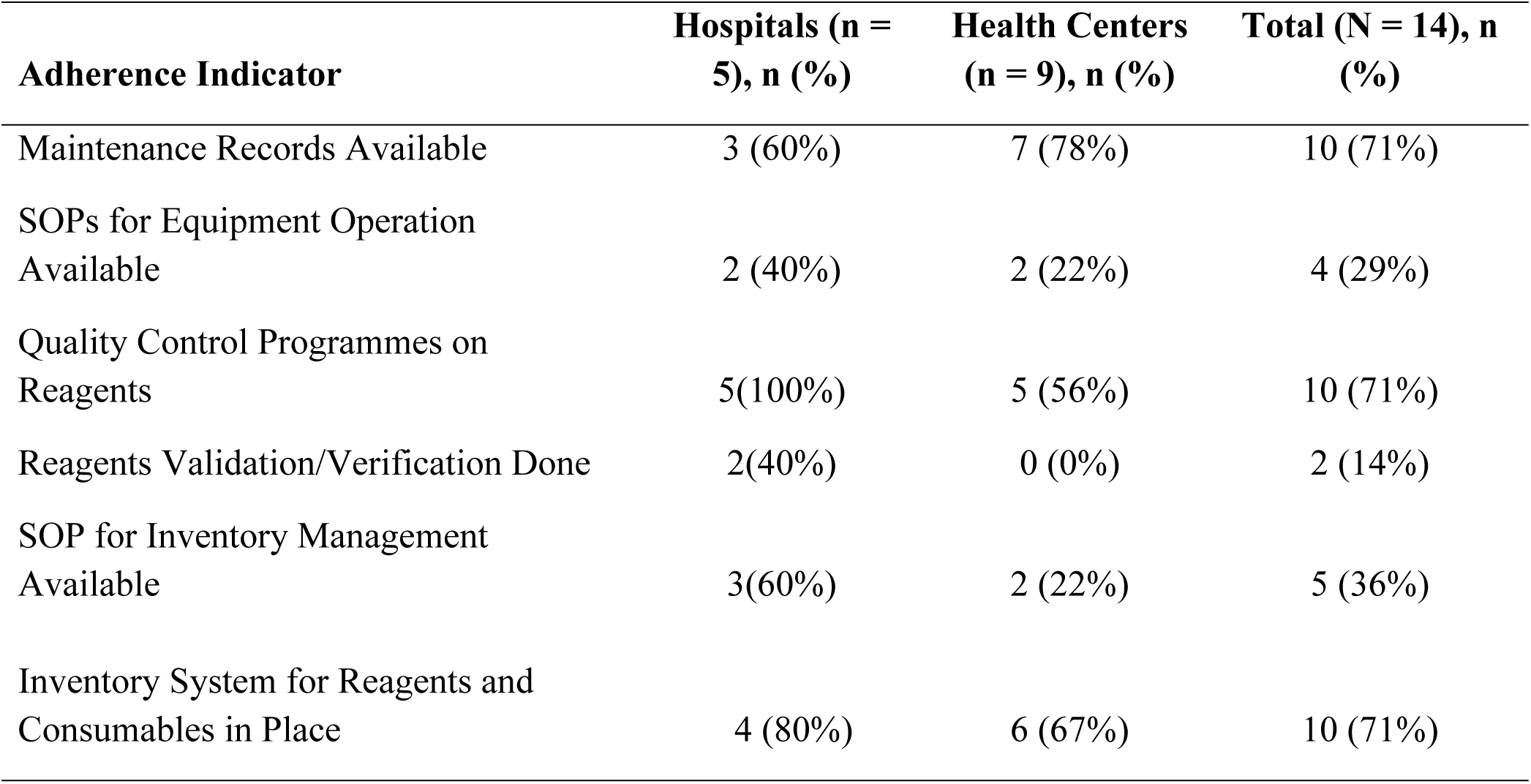
Adherence to Equipment, Consumable and Reagents Standards (N=14)

### Challenges in STI Testing Reported by Healthcare Facilities

The healthcare facilities reported several challenges in STI testing, with the most common being the lack of testing guidelines (85.7%) and standard operating procedures (71.4%). Other significant issues included reagent shortages (64.3%) and the absence of on-the-job training programs (57.1%) (Table 4).

**Table 4:**
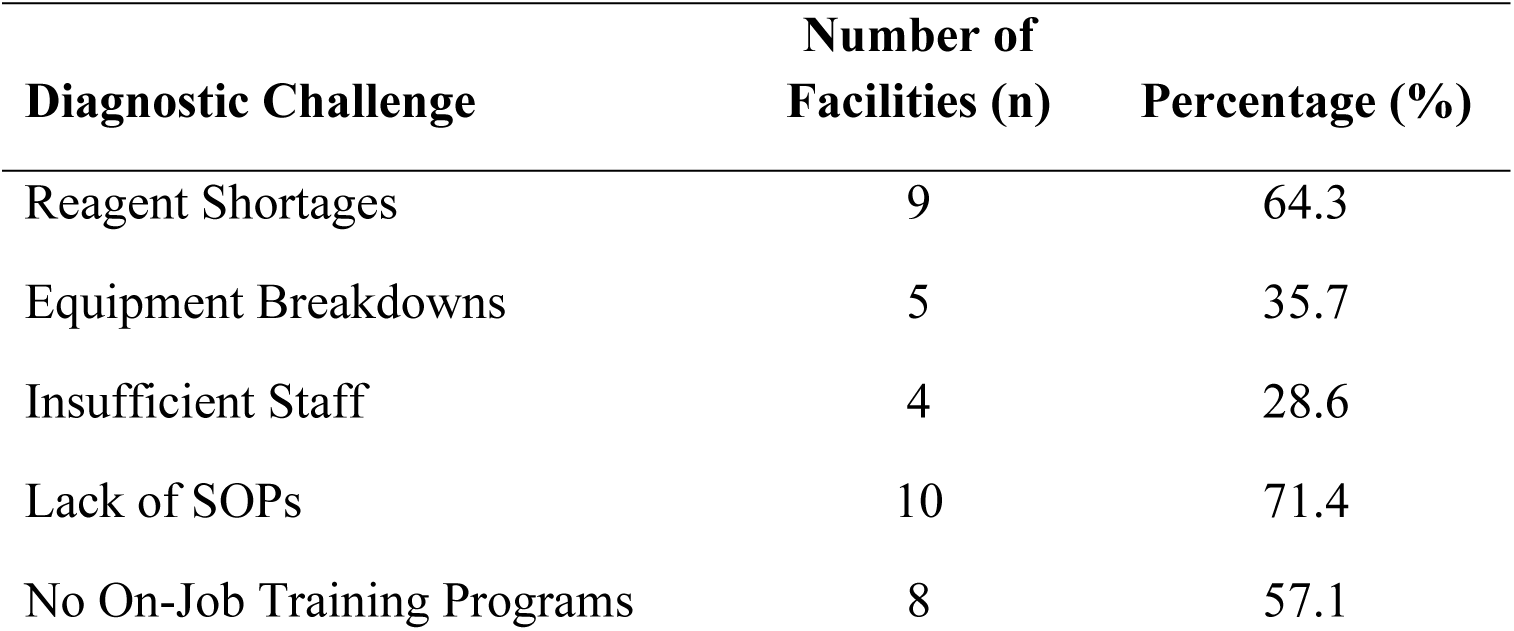
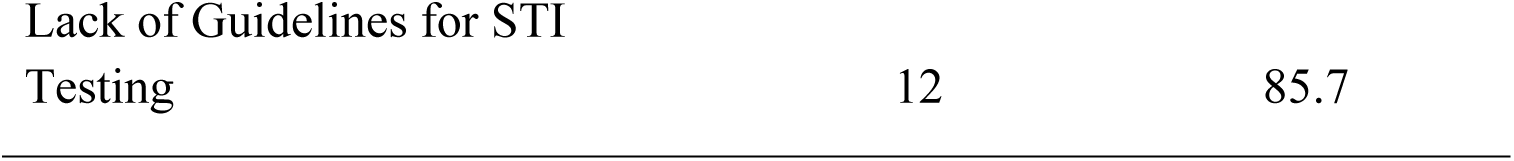
Challenges in STI Diagnosis Reported by Healthcare facilities (N=14)

## Discussion

The global burden of curable STIs is rising, with low resource settings disproportionately affected. Combined with the escalation of AMR in key STI pathogens, this creates an urgent need to strengthen diagnostic capacity and control efforts in order to effectively curb ongoing transmission (21). This study provides key insights into the laboratory capacity for STI testing within Moshi Municipality, Tanzania. The STI tests performed at health facilities were evaluated against national standards for medical laboratory requirements. Findings indicate that laboratories in Moshi require substantial improvement and monitoring to meet these standards for STI testing. This enhanced need applies to all assessed facilities, including health centers and hospitals. Although data presented here are from Moshi only, the observed gaps are likely to be indicative of broader challenges affecting laboratory services for STI diagnosis across Tanzania as a whole.

These local findings are consistent with national evidence showing that the quality and capacity of laboratory services at primary healthcare (PHC) level in Tanzania remain low, with a critical shortage of qualified laboratory personnel, particularly in public facilities serving rural populations. Even among laboratories that are operational, more than half do not perform internal quality control or participate in external quality assessments (EQA), raising concerns on the reliability and comparability of test results (22).

Reassuringly, most health facilities in this study were found to undertake routine rapid diagnostic tests (RDTs) for HIV, syphilis, and hepatitis B, given the global and national focus on eliminating mother-to-child transmission of these infections under the WHO’s triple elimination initiative (23).

This initiative promotes integrated testing and care for HIV, syphilis, and hepatitis B in antenatal services to reduce vertical transmission and improve maternal and child health outcomes (23). In Tanzania, HIV and syphilis testing have become routine components of antenatal care, supported by national policies and widespread use of rapid test kits such as the SD Bioline HIV/syphilis duo (SD Standard Diagnostic, INC, Korea) provided free of charge in public and private facilities (24). In parallel, national HIV prevention programmes are also expanding the use of oral pre-exposure prophylaxis (PrEP) within broader combination prevention strategies for key and priority populations. PrEP is specifically offered to sex workers, men at high risk, people who inject drugs, vulnerable adolescent girls (15–19 years), young women (20–24 years), high-risk pregnant and breastfeeding women, and HIV negative partners in serodiscordant relationships where the partner living with HIV has not yet achieved viral suppression below 50 copies/ml (25). However, despite the inclusion of hepatitis B in the triple elimination strategy, hepatitis B screening has not yet been consistently integrated into routine antenatal care services. Implementation has only recently begun in a limited number of healthcare facilities, starting in the last quarter of 2025, and nationwide coverage remains incomplete.

Recent national surveillance data indicate that syphilis remains a persistent problem among pregnant women in Tanzania, with an overall prevalence of about 1.4% and marked regional variation, reaching over 4% in some regions (26). Given the substantial risk of adverse pregnancy outcomes associated with untreated maternal syphilis, this continuing burden underscores the importance of reliable syphilis screening, confirmation, and treatment within antenatal and general STI services (26). Consistent with WHO guidance, Tanzania’s preventing mother-to-child transmission (PMTCT) programme recommends lifelong triple ART (Option B+) for all pregnant and breastfeeding women living with HIV regardless of CD4 count or clinical stage, together with antiretroviral prophylaxis and early virological testing for HIV-exposed infants to minimise mother-to-child transmission (27). The combined efforts under the WHO triple elimination framework have contributed to improved HIV testing coverage and the expansion of syphilis testing within antenatal care services, particularly in high-burden countries such as those in sub-Saharan Africa (28).

The National Standard for Medical Laboratories specifies that RDTs should be available in healthcare facilities. However, it does not define which specific RDTs should be used despite substantial differences in sensitivity and specificity. In our study, all facilities reported using RDTs from various manufacturers and indicated that procurement decisions were driven primarily by availability and cost rather than by documented performance characteristics. A major concern highlighted by this study is the severe lack of diagnostic capacity for CT and Ng, the most prevalent bacterial STIs globally and in our region (2, 13). Although national guidelines recommend at least point-of-care testing (POCTs) for CT, none of the 14 health facilities included in this study performed any testing for this pathogen. POCTs for STIs have the potential to improve access to diagnostics in LMICs, support more accurate identification of causative pathogens, and enable timely treatment, which in turn reduces overtreatment, transmission, and long-term health complications. POCTs may ultimately replace the syndromic approach commonly used in Tanzania and elsewhere in LMICs.

The WHO introduced the ASSURED framework, later expanded to REASSURED, to guide key features that POCTs should have for their use in LMICs. These include real-time data connectivity, easy specimen collection, affordability, high sensitivity and specificity, user friendliness, rapid and robust performance, minimal equipment requirements, and wide accessibility to end users (29).

Several nucleic acid amplification based POCTs for chlamydia and gonorrhoea are currently available and are being used mainly in high-income settings and in some LMICs with donor support. This includes the binx io CT/GC test (binx health, Boston, MA U.S.A.), the Xpert CT/NG assay on the GeneXpert platform (Cepheid, Sunnyvale, California, US) and the Visby Sexual Health test (Visby Health, San Jose, CA U.S.A.). However, these platforms do not fully meet the WHO REASSURED criteria, largely because of their high cost and stringent reagent storage requirements, which limits their suitability for routine use in resource-limited settings (30). Apart from costs, there is no reliable or sustainable market for POCTs to detect STI antibiotic resistance in LMICs (31). Tanzania currently has more than 300 GeneXpert instruments deployed mainly across zonal, regional, district hospitals and selected high-volume health centres primarily used for tuberculosis diagnosis (32,33). Existing diagnostic platforms could potentially be leveraged and expanded to support STI testing. However, financial constraints continue to limit such integration in LMICs contexts. There is an urgent need therefore for new POCTs that are fast, reliable, and affordable to help address the growing global burden of STIs and AMR.

Treatment of gonorrhoea, along with widespread over the counter, prescription free antibiotic use in the community, inevitably exerts selective pressure on *Ng* and drives the emergence of AMR. Bacterial culture remains a cornerstone for tracking AMR, because it allows laboratories to detect new resistant strains and keep treatment guidelines aligned with local susceptibility patterns. In this study, three hospitals reported having microbiology laboratories capable of culturing specimens from patients with suspected gonorrhoea. However, none reported recovering Ng isolates, suggesting that, despite nominal culture capacity, functional recovery of Ng is not achieved in routine practice. This finding reflects a broader challenge observed across many African countries, where insufficient culture based surveillance limits the ability to monitor AMR trends and detect emerging resistance in Ng (34). This challenge extends beyond gonococcal culture to bacteriology and antimicrobial susceptibility testing (AST) more broadly, where limited adherence to national and international standards and best laboratory practices remains a common issue. Evidence from a multi-country assessment across 14 sub-Saharan African countries highlights this gap, showing that among more than 53,000 documented clinical laboratories, only about 1% were formally designated to conduct bacteriological testing, with an even smaller proportion having the capacity to perform AST (35). From a technical perspective, culturing Ng and performing phenotypic AST is challenging. The organism is fastidious, requiring enriched media and incubation in a carbon dioxide enriched atmosphere, and it is highly sensitive to environmental conditions, including delays or suboptimal conditions during specimen collection, transport, and processing (36). Even the choice of swab can be critical; wooden shafts and cotton tips can inhibit or kill Ng, whereas plastic or wire shafts with rayon, dacron, or calcium alginate tips are recommended for culture (37). To bridge the gap between nominal and functional capacity, laboratories should routinely use Ng reference strains to verify media, incubation conditions and technical procedures, combined with targeted training and mentorship for staff in gonococcal culture and AST. Several factors likely contribute to the low recovery of Ng, including limited technical expertise and inconsistent availability of consumables and reagents. Addressing these gaps will require sustained investment in laboratory infrastructure, training, and quality systems. Strengthening culture and AST capacity is urgently needed to improve diagnostic capability and keep pace with the rapidly evolving threat of antibiotic resistance in the region.

Whole genome sequencing (WGS) has emerged as a powerful tool for advancing our understanding of infectious pathogens by enabling detailed analyses of their transmission dynamics, evolutionary relationships, and the genetic determinants of AMR. However, despite the critical role of genomic data in informing STI transmission and resistance, most sequencing studies continue to originate from high income countries, with limited representation from LMIC settings such as Tanzania (38). This gap is particularly evident for Ng, where genomic data are essential for tracking the emergence and spread of resistance to first line antibiotics, yet remain scarce across much of sub-Saharan Africa (38). When pathogen genomic data are integrated with detailed demographic and behavioural information, they provide a high resolution view of how infections circulate within and between key populations. The recent multinational mpox outbreak, largely driven by dense sexual networks and rapid international travel, illustrates how quickly emerging pathogens can spread and highlights the importance of timely genomic surveillance (39–41).

## Conclusion

Substantial investment is needed to strengthen laboratory capacity for the aetiological diagnosis of STIs in Tanzania, particularly for Ng and other key pathogens. This includes sustained support for infrastructure, equipment, supplies, and human resources, as well as robust quality systems to ensure reliable culture, antimicrobial susceptibility testing, and molecular diagnostics. Doxycycline post-exposure prophylaxis (DoxyPEP), which has been shown to reduce CT and syphilis and is already being deployed in several countries, could eventually be beneficial in the Tanzanian context. However, without adequate laboratory infrastructure to monitor STI incidence and resistance, the impact and safety of such preventive strategies cannot be properly assessed. Similarly, emerging vaccines such as meningococcal B vaccine (4CMenB) with partial protection against gonorrhoea and other OMV based or protein based gonococcal candidates alongside vaccine candidates for chlamydia and herpes simplex virus (HSV-2), are being evaluated mainly in high-income settings. Understanding the locally circulating strains and resistance patterns will be crucial both for guiding the introduction and evaluation of these new interventions and for improving antibiotic stewardship.

## Data Availability

The minimal data set [and accompanying code, where generated] is available at Figshare via10.6084/m9.figshare.32228730

http://10.6084/m9.figshare.32228730

## Abbreviations

AMR: Antimicrobial Resistance
ART: Antiretroviral Therapy
AST: Antimicrobial Susceptibility Testing
CT: Chlamydia trachomatis
EQA: External Quality Assessment
HCs: Health Centers
HPV: Human Papillomavirus
HSV-2: Herpes Simplex Virus-2
IQR: Interquartile Range
KCMC: Kilimanjaro Christian Medical Centre
LMICs: Low– and Middle-Income Countries
MG: Mycoplasma genitalium
NASHCoP: National AIDS, STIs and Hepatitis Control Programme
Ng: Neisseria gonorrhoeae
PHC: Primary Healthcare
PMTCT: Preventing Mother-to-Child Transmission
POC(T): Point-of-Care (Test)
PrEP: Pre-Exposure Prophylaxis
RDT(s): Rapid Diagnostic Test(s)
SOP(s): Standard Operating Procedure(s)|
STI(s): Sexually Transmitted Infection(s)
TP(HA): Treponema pallidum (Hemagglutination Assay)
TPPA: Treponema pallidum Particle Agglutination Assay
TV: Trichomonas vaginalis|
WHO: World Health Organization
WGS: Whole Genome Sequencing

## Acknowledgements

We express our sincere gratitude to all participating healthcare facilities and healthcare workers in Moshi Municipal Council for their invaluable support and cooperation during data collection.

## Supporting information

**S1 Fig. Map showing the locations of participating healthcare facilities in Moshi Municipal Council**.

